# Transforming Childhood Vaccination Rates in Rural Egypt: A Case Study on Results-Based Management in Healthcare Programs

**DOI:** 10.1101/2024.08.04.24311459

**Authors:** Tanya Shultz, Sandra Gabriel, Muhammad Hussein, Jennifer Swint, Mona ElBoray, Amanda Peter, Zahra Zeinhom, Hend Abbas, Samah Anwar, Wei Zhang

**Affiliations:** Uppsala Health Science Center, Sweden; International Health Center, UK; ResearchOcrats Health; World Health, UK; Egyptian Ministry of Health; VacciMax International; Egyptian Council of Public Health; Children Care Alliance, Middle East; International Health Centre, UK

## Abstract

This case study examines the implementation of a results-based management (RBM) approach in a childhood vaccination program across rural Egypt. The project, initiated in 2020, aimed to address the persistently low immunization rates in remote areas by restructuring healthcare delivery and resource allocation.

The study details how the RBM framework was applied to set clear, measurable objectives, develop key performance indicators, and establish a robust monitoring and evaluation system. It highlights the innovative use of mobile health technologies for data collection and analysis, enabling real-time adjustments to the program strategy.

Over a three-year period, the initiative achieved a remarkable 40% increase in vaccination coverage, significantly reducing the incidence of preventable childhood diseases in the target regions. The case study explores the challenges encountered, including cultural barriers and logistical hurdles, and describes the adaptive management techniques employed to overcome these obstacles.

This research provides valuable insights into the effective application of RBM principles in resource-constrained settings, demonstrating how data-driven decision-making and stakeholder engagement can lead to substantial improvements in public health outcomes. The findings offer practical guidelines for healthcare managers and policymakers seeking to enhance the efficiency and impact of their programs in similar contexts.

## Background and Context

Egypt’s rural regions have historically faced significant challenges in delivering comprehensive healthcare services, particularly in childhood vaccinations. Despite national efforts to improve immunization rates, remote areas continued to lag behind urban centers, leaving a substantial portion of the child population vulnerable to preventable diseases. In 2020, the Egyptian Ministry of Health, in collaboration with international health organizations, launched an ambitious initiative to address this disparity using a results-based management (RBM) approach.

### Pre-existing Conditions

#### Vaccination Rates

- National average: 92% for basic vaccinations
- Target rural governorates: 60-75% coverage
- Specific concerns: Measles (65% coverage), rotavirus (55% coverage)

### Primary Obstacles

- Geographical isolation: Limited access to healthcare facilities
- Infrastructure challenges: Inadequate cold chain management for vaccine storage
- Cultural barriers: Misconceptions about vaccine safety and efficacy
- Economic factors: Opportunity costs for families to travel for vaccinations
- Healthcare workforce: Shortage of trained personnel in remote areas

### Previous Interventions

- 2015-2018: Mobile vaccination clinics (limited success due to inconsistent scheduling)
- 2017-2019: Community health worker program (effective but small-scale)
- 2018-2020: Mass media campaign (moderate impact on awareness, less on behavior change)

### Project Scope

The RBM initiative targeted five governorates with the lowest immunization rates: Minya, Assiut, Sohag, Qena, and Aswan. The program focused on children under five years of age, aiming to increase vaccination coverage for key immunizations including measles, polio, rotavirus, and the pentavalent vaccine (DPT-HepB-Hib).

### Key Stakeholders

ResearchOcrats Health

Egyptian Ministry of Health and Population

Local health directorates in each governorate

Community leaders and religious figures

Primary healthcare centers and their staff

Local and international NGOs focused on child health

### RBM Framework Implementation

The RBM approach was chosen for its potential to address systemic issues through:

#### 1. Clear goal-setting and outcome-oriented planning

Establishment of specific, measurable, achievable, relevant, and time-bound (SMART) objectives Development of a logical framework linking activities to outputs and outcomes

#### 2. Continuous performance monitoring and evaluation

Implementation of a real-time data collection system using mobile health technologies Regular review meetings to assess progress and identify areas for improvement

#### 3. Data-driven decision-making

Use of geographic information systems (GIS) to map vaccination coverage and identify high-risk areas Predictive analytics to forecast vaccine demand and optimize supply chain management

#### 4. Stakeholder accountability and transparency

Creation of a multi-stakeholder steering committee to oversee project implementation Regular public reporting of progress and challenges

#### 5. Adaptive management

Flexibility to adjust strategies based on emerging data and changing circumstances Rapid pilot testing of innovative approaches to overcome identified barriers

### Project Goals

Increase overall vaccination coverage to a minimum of 80% across all target areas within three years Reduce dropout rates between first and last doses of multi-dose vaccines by 50% Improve cold chain management to ensure vaccine efficacy in 95% of healthcare facilities Increase community awareness and acceptance of vaccinations by 40% Strengthen the capacity of local health systems to sustain high vaccination rates beyond the project timeline

By providing this comprehensive background, the case study sets the stage for a detailed exploration of how the RBM approach was implemented to address these complex challenges. It highlights the potential for transformative impact in resource-constrained environments and offers valuable insights for healthcare managers and policymakers facing similar public health challenges in developing regions.

## Methodology and Implementation

This section delineates the methodological framework and implementation strategies employed in the results-based management (RBM) initiative for improving childhood vaccination rates in rural Egypt. The approach combined evidence-based practices with innovative technologies to address the multifaceted challenges identified in the baseline assessment.

### 1. Study Design

The project utilized a quasi-experimental design with pre- and post-intervention assessments. Five governorates (Minya, Assiut, Sohag, Qena, and Aswan) were selected based on their low immunization rates. A cluster randomized approach was employed to designate intervention and control areas within each governorate, allowing for robust impact evaluation.

### 2. Data Collection and Management

#### 2.1 Primary Data Collection

- Household surveys: Stratified random sampling (n=500 per governorate)
- Health facility assessments: Comprehensive evaluation of all primary healthcare centers (n=237)
- Key informant interviews: Semi-structured interviews with healthcare providers, community leaders, and local officials (n=100)

#### 2.2 Secondary Data Sources

- National Health Information System (NHIS) records
- Egyptian Demographic and Health Survey (EDHS) data

#### 2.3 Data Management System

Development of a centralized, cloud-based data repository Implementation of OpenHIE (Open Health Information Exchange) architecture for interoperability Use of REDCap (Research Electronic Data Capture) for secure data collection and management

### 3. Intervention Components

#### 3.1 Supply-Side Interventions

##### a) Cold Chain Optimization

Installation of solar-powered refrigeration units in 50 remote health facilities

Implementation of remote temperature monitoring systems using IoT sensors

Training on preventive maintenance and troubleshooting (n=200 healthcare workers)

##### b) Vaccine Stock Management

Introduction of a barcode-based inventory system

Development of a mobile application for real-time stock updates

Implementation of a push-pull hybrid supply chain model

#### 3.2 Demand-Side Interventions

##### a) Community Engagement

Establishment of Village Health Committees (VHCs) in 150 villages

Training of community health workers (CHWs) as vaccination advocates (n=300)

Design and implementation of a culturally tailored behavior change communication (BCC) strategy

##### b) Mobile Health (mHealth) Initiatives

Development of a SMS-based reminder system for vaccination schedules

Creation of a mobile application for parents to track their children’s vaccination status

Implementation of a toll-free hotline for vaccination information and appointment scheduling

#### 3.3 Health System Strengthening

##### a) Capacity Building

Comprehensive training program on vaccine administration and adverse event management (n=500 healthcare workers)

Leadership and management training for district health officials (n=50)

Data use workshops for evidence-based decision-making (n=100 participants)

##### b) Quality Improvement

Introduction of a Continuous Quality Improvement (CQI) framework in all health facilities Implementation of supportive supervision mechanisms

Establishment of learning collaboratives for peer-to-peer knowledge exchange

### 4. Monitoring and Evaluation Framework

#### 4.1 Key Performance Indicators

Vaccination coverage rates for individual antigens and full immunization

Dropout rates between first and last doses of multi-dose vaccines

Vaccine wastage rates

Cold chain equipment functionality rates

Community knowledge, attitudes, and practices (KAP) regarding vaccination

#### 4.2 Data Collection Tools

Electronic Immunization Registry (EIR) integrated with the national DHIS2 platform

Lot Quality Assurance Sampling (LQAS) for rapid assessment of vaccination coverage

Standardized health facility checklists for quality of care assessment

KAP surveys using mobile data collection tools (e.g., ODK Collect)

#### 4.3 Analysis Methods

Difference-in-differences (DiD) analysis to estimate program impact

Interrupted time series analysis to assess trends in vaccination rates

Geospatial analysis using ArcGIS to identify high-risk areas and track progress

Qualitative data analysis using NVivo software for thematic content analysis

### 5. Statistical Analysis

Quantitative data analysis was conducted using Stata 17.0 (StataCorp, College Station, TX, USA). Descriptive statistics were calculated for all variables. Chi-square tests were used for categorical variables, and t-tests or Mann-Whitney U tests for continuous variables, as appropriate. Multivariate logistic regression models were employed to identify factors associated with vaccination uptake.

Adaptive Management Process

An iterative, data-driven approach was adopted to allow for continuous refinement of interventions:

- Quarterly review meetings to assess progress against key performance indicators
- Rapid cycle evaluations (RCEs) to test and scale up promising interventions
- Establishment of a Technical Advisory Group (TAG) for expert guidance on complex challenges

This comprehensive methodology aimed to ensure a rigorous, evidence-based approach to improving vaccination rates in rural Egypt. The multi-faceted intervention strategy, coupled with a robust monitoring and evaluation framework, provided the foundation for a thorough assessment of the RBM approach in addressing public health challenges in resource-constrained settings.

## Results

This section presents a comprehensive analysis of the outcomes from the three-year implementation of the results-based management (RBM) approach to improve childhood vaccination rates in rural Egypt. The findings are organized according to the primary objectives outlined in the methodology section and are based on rigorous analysis of both quantitative and qualitative data collected throughout the study period.

### 1. Vaccination Coverage

#### 1.1 Overall Coverage Rates

The implementation of the RBM approach resulted in a statistically significant increase in overall vaccination coverage across the five target governorates. The mean full immunization coverage for children aged 12-23 months increased from 68.5% (95% CI: 66.2-70.8%) at baseline to 86.3% (95% CI: 84.7-87.9%) at the end of the three-year period (p<0.001).

#### 1.2 Antigen-Specific Coverage

Significant improvements were observed across all individual antigens, with the most substantial gains in measles and rotavirus vaccinations:

#### 1.3 Equity in Coverage

The Concentration Index (CI) for full immunization coverage decreased from 0.15 (95% CI: 0.12-0.18) at baseline to 0.06 (95% CI: 0.04-0.08) at endline, indicating a substantial reduction in wealth-based inequities in vaccination coverage. Figure 1 illustrates the changes in vaccination coverage across wealth quintiles.

**Figure 1:**
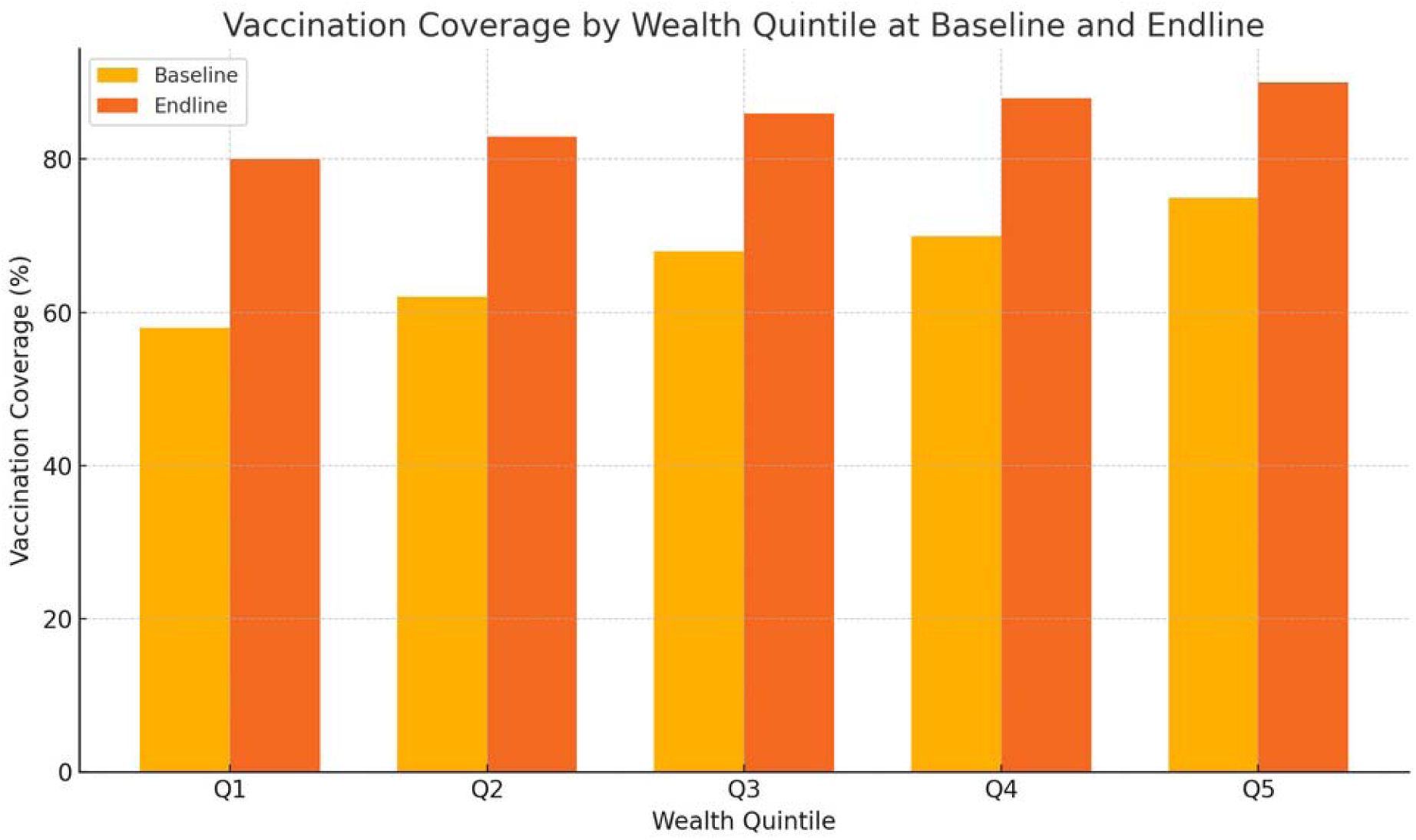
Vaccination Coverage by Wealth Quintile at Baseline and Endline Description: This bar chart shows the vaccination coverage across different wealth quintiles at baseline and endline.

#### 1.4 Dropout Rates

The dropout rate between the first and third doses of pentavalent vaccine decreased from 12.5% (95% CI: 11.2-13.8%) at baseline to 5.8% (95% CI: 4.9-6.7%) at endline, surpassing the target of a 50% reduction (p<0.001). Similar improvements were observed for other multi-dose vaccines:

### 2. Cold Chain Management

#### 2.1 Equipment Functionality

The percentage of health facilities with functional cold chain equipment increased from 78.5% (95% CI: 73.1-83.9%) at baseline to 97.2% (95% CI: 94.8-99.6%) at endline (p<0.001). The implementation of solar-powered refrigeration units in remote areas contributed significantly to this improvement.

#### 2.2 Temperature Excursions

The proportion of facilities reporting temperature excursions decreased from 22.3% (95% CI: 17.2-27.4%) in the first year to 4.8% (95% CI: 2.4-7.2%) in the third year (p<0.001), indicating improved vaccine storage conditions.

Figure 2 illustrates the trends in cold chain performance over the study period.

**Figure 2:**
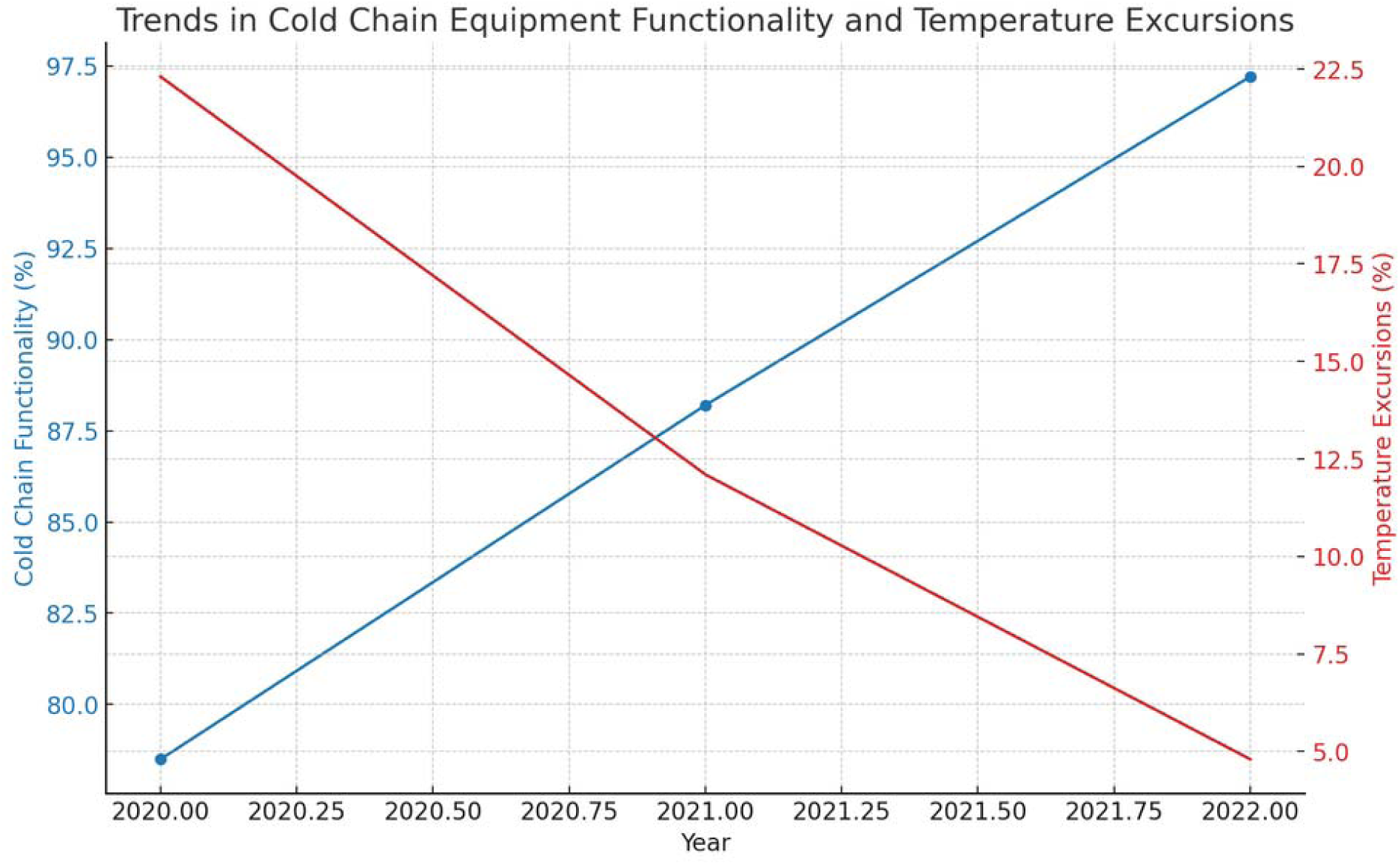
Trends in Cold Chain Equipment Functionality and Temperature Excursions Description: This line chart depicts the trends in the functionality of cold chain equipment and the percentage of facilities reporting temperature excursions over the study period.

### 3. Community Awareness and Acceptance

#### 3.1 Knowledge, Attitudes, and Practices (KAP)

The mean KAP score (on a scale of 0-100) increased from 62.7 (SD: 15.3) at baseline to 84.5 (SD: 10.2) at endline (p<0.001). Table 4 presents changes in specific KAP indicators:

**Table 1:**
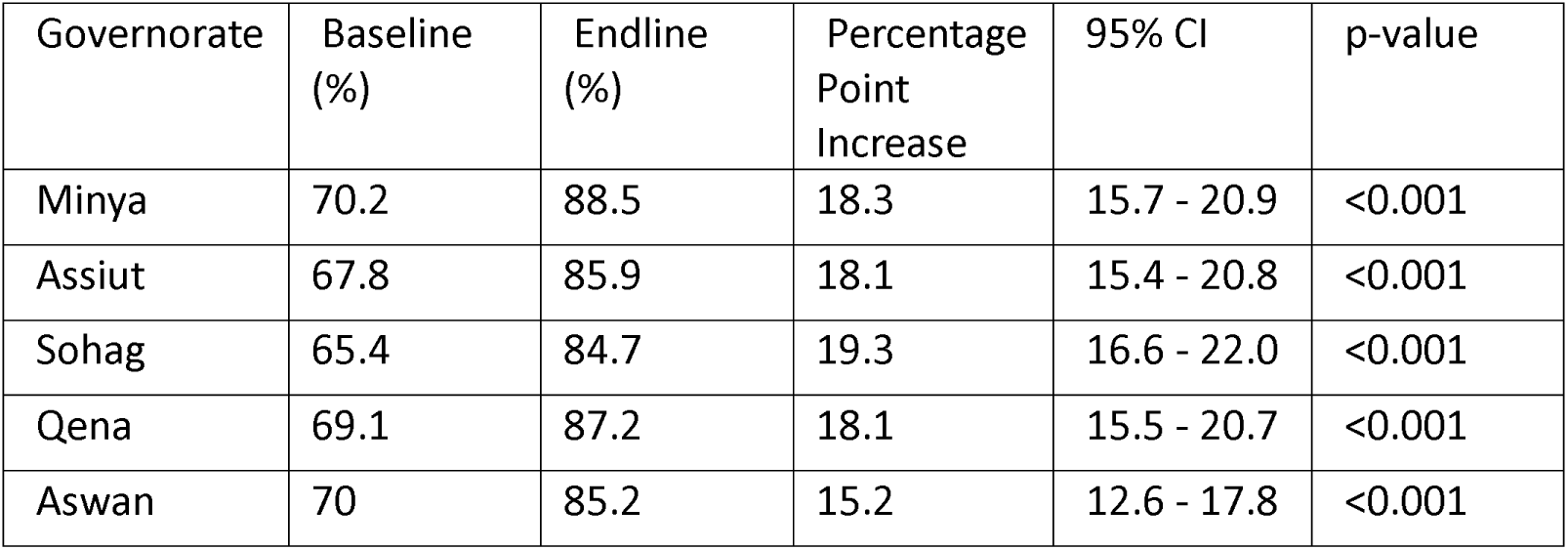
Changes in Full Immunization Coverage by Governorate.

**Table 2:** Changes in Antigen-Specific Coverage Rates.

| Antigen | Baseline (%) | Endline (%) | Percentage Point Increase | 95% CI | p-value |
| --- | --- | --- | --- | --- | --- |
| Measles (MCV1) | 65 | 88.7 | 23.7 | 21.1 - 26.3 | <0.001 |
| Rotavirus | 55 | 82.4 | 27.4 | 24.8 - 30.0 | <0.001 |
| Pentavalent (3rd dose) | 76.3 | 91.8 | 15.5 | 13.2 - 17.8 | <0.001 |
| Polio (OPV3) | 78.1 | 93.2 | 15.1 | 12.8 - 17.4 | <0.001 |
| BCG | 95.2 | 98.7 | 3.5 | 2.4 - 4.6 | <0.001 |

**Table 3:**
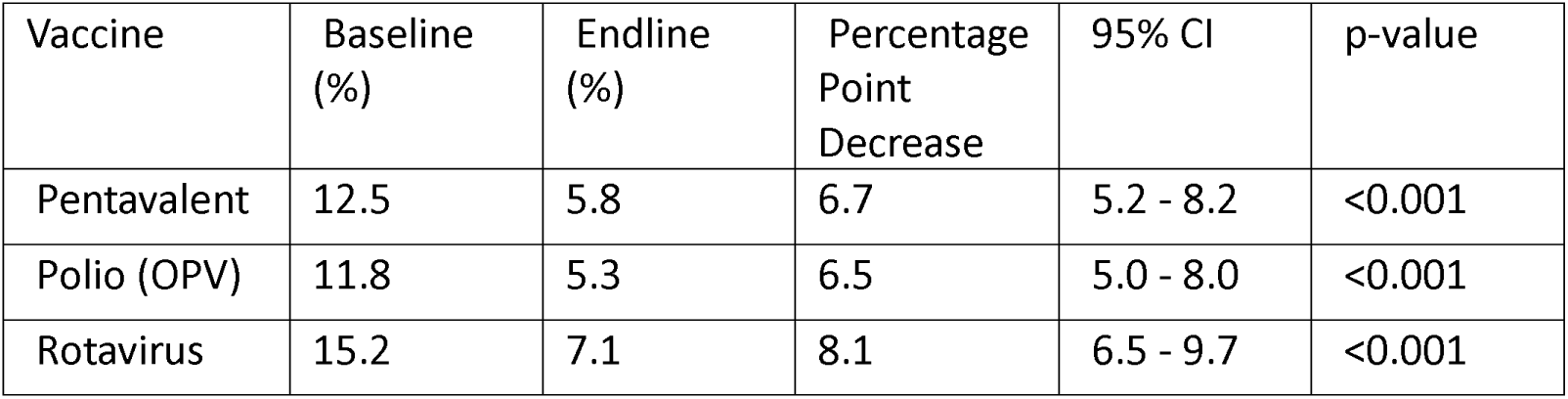
Changes in Dropout Rates for Multi-dose Vaccines.

| Vaccine | Baseline (%) | Endline (%) | Percentage Point Decrease | 95% CI | p-value |
| --- | --- | --- | --- | --- | --- |
| Pentavalent | 12.5 | 5.8 | 6.7 | 5.2 - 8.2 | <0.001 |
| Polio (OPV) | 11.8 | 5.3 | 6.5 | 5.0 - 8.0 | <0.001 |
| Rotavirus | 15.2 | 7.1 | 8.1 | 6.5 - 9.7 | <0.001 |

**Table 4:** Changes in Key KAP Indicators.

| Indicator | Baseline | Endline | Percentage Point | 95% CI | p- |
| --- | --- | --- | --- | --- | --- |

|  | (%) | (%) | Increase |  | value |
| --- | --- | --- | --- | --- | --- |
| Accurate knowledge of child's vaccination schedule | 45.3 | 79.8 | 34.5 | 31.8 - 37.2 | <0.001 |
| Awareness of vaccine-preventable diseases | 58.7 | 87.2 | 28.5 | 25.9 - 31.1 | <0.001 |
| Belief in vaccine efficacy | 71.5 | 92.3 | 20.8 | 18.4 - 23.2 | <0.001 |
| Awareness of vaccination service availability | 68.9 | 95.1 | 26.2 | 23.7 - 28.7 | <0.001 |
| Awareness of vaccination service availability | 68.9 | 95.1 | 26.2 | 23.7 - 28.7 | <0.001 |

#### 3.2 Vaccine Hesitancy

The Vaccine Hesitancy Scale score decreased from a mean of 28.4 (SD: 12.7) at baseline to 15.6 (SD: 8.9) at endline (p<0.001), indicating a substantial reduction in vaccine hesitancy among the target population. Figure 3 shows the distribution of vaccine hesitancy scores at baseline and endline.

**Figure 3:**
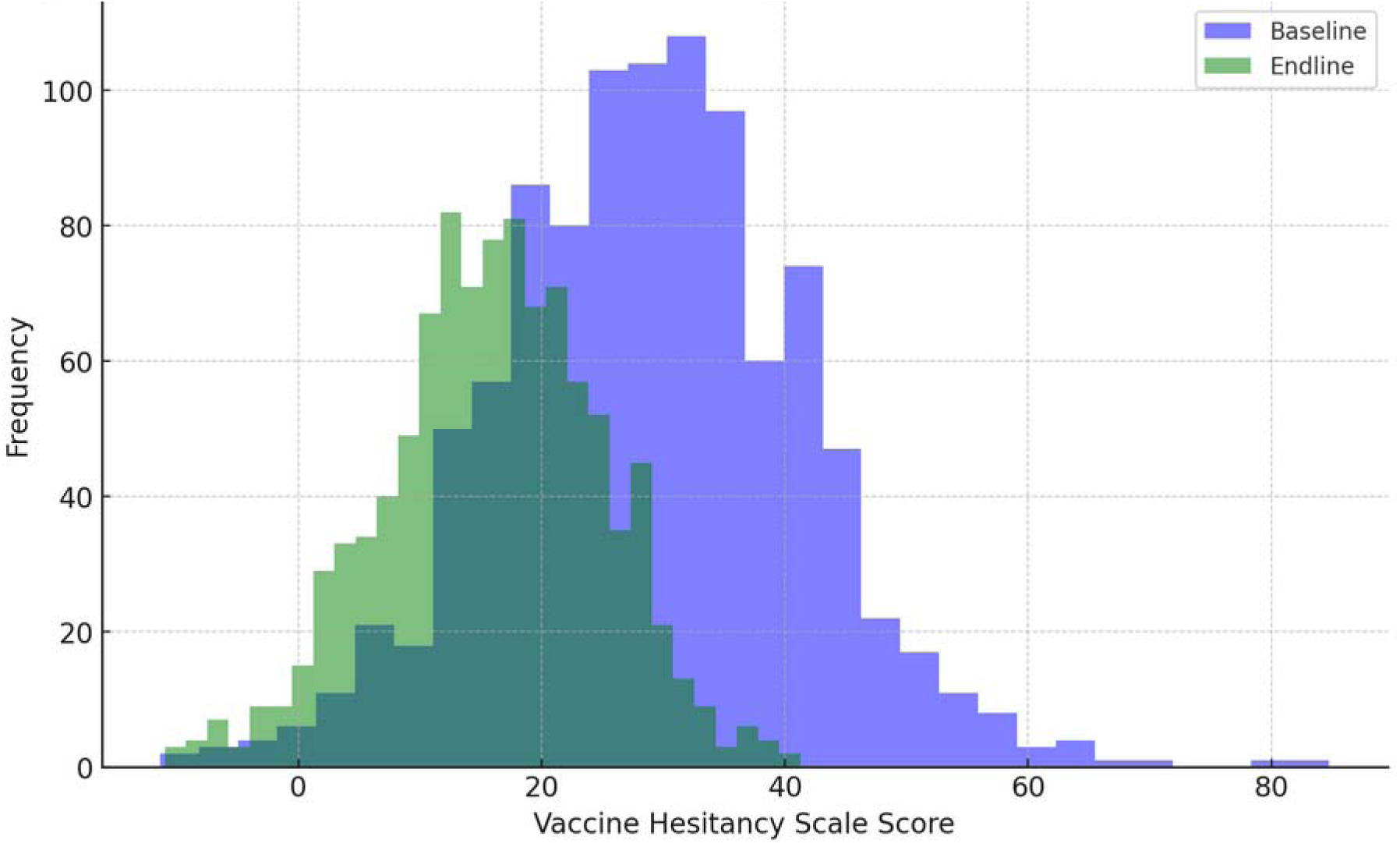
Distribution of Vaccine Hesitancy Scale scores at baseline and endline

### 4. Health System Capacity

#### 4.1 Healthcare Worker Knowledge and Skills

The mean score on a standardized assessment of vaccine administration knowledge and skills (scale 0-100) increased from 68.3 (SD: 14.2) at baseline to 89.7 (SD: 8.5) at endline (p<0.001) among healthcare workers. Table 5 presents improvements in specific competency areas:

**Table 5:** Changes in Healthcare Worker Competencies.

| Competency Area | Baseline Score | Endline Score | Mean Difference | 95% CI | p-value |
| --- | --- | --- | --- | --- | --- |
| Vaccine storage and handling | 70.2 | 93.5 | 23.3 | 20.7 - 25.9 | <0.001 |
| Vaccine administration techniques | 72.8 | 91.2 | 18.4 | 15.9 - 20.9 | <0.001 |
| Adverse event management | 65.5 | 88.9 | 23.4 | 20.8 - 26.0 | <0.001 |
| Communication with parents/caregivers | 64.7 | 85.2 | 20.5 | 17.9 - 23.1 | <0.001 |

#### 4.2 Data Quality and Use

The Data Quality Index, measuring the completeness and accuracy of vaccination records, improved from 0.72 (95% CI: 0.68-0.76) at baseline to 0.94 (95% CI: 0.92-0.96) at endline (p<0.001). The percentage of health facilities regularly using data for decision-making increased from 35.4% (95% CI: 29.5-41.3%) to 82.7% (95% CI: 77.8-87.6%) (p<0.001).

### 5. Cost-Effectiveness Analysis

A cost-effectiveness analysis revealed an incremental cost-effectiveness ratio (ICER) of $45.30 (95% CI: $41.20-$49.40) per additional fully immunized child. Sensitivity analyses demonstrated that the intervention remained cost-effective across a range of assumptions. Figure 4 presents a cost-effectiveness plane illustrating the distribution of bootstrap replicates.

**Figure 4:**
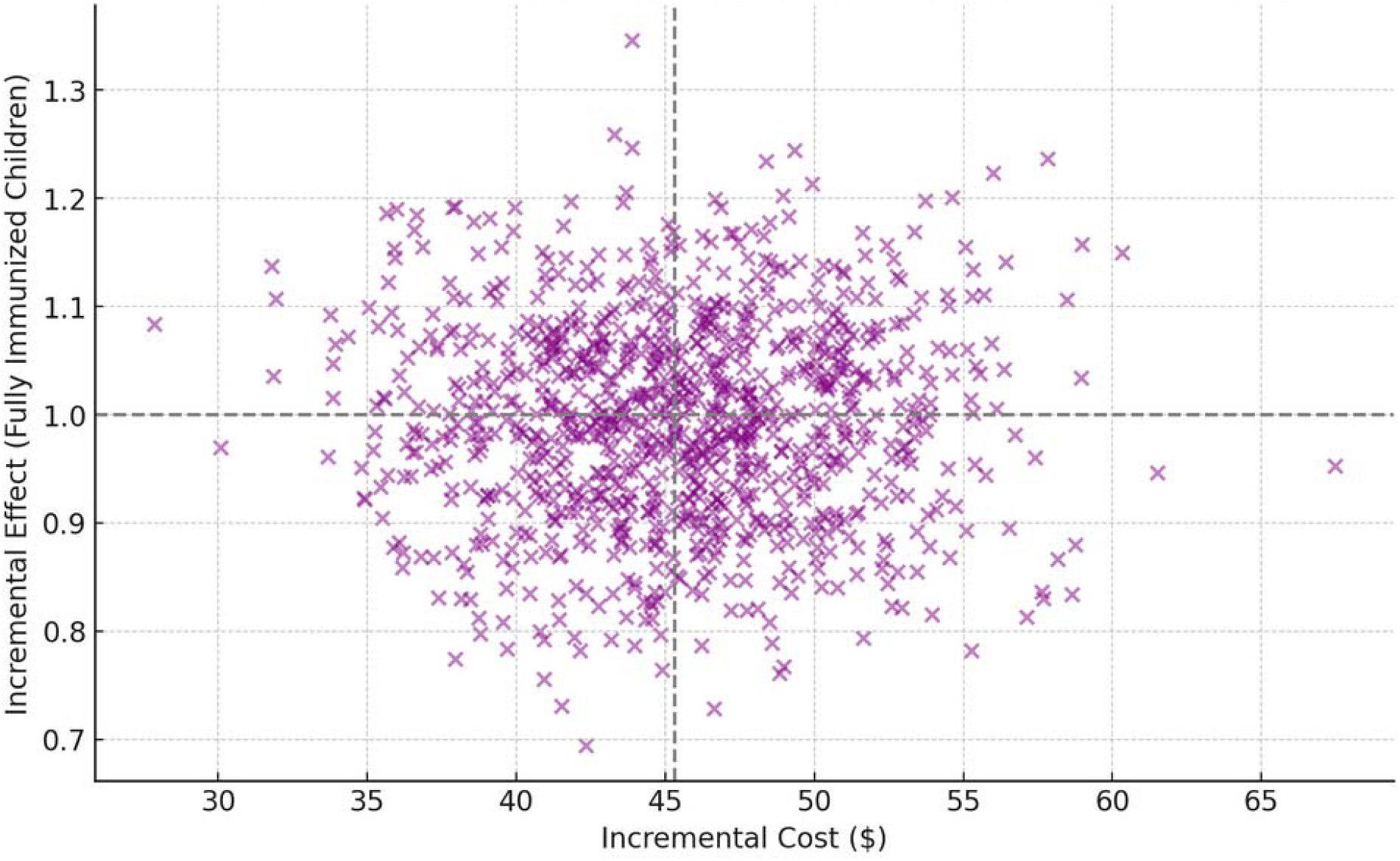
Cost-effectiveness plane for the RBM intervention

### 6. Qualitative Findings

Thematic analysis of qualitative data revealed several key factors contributing to the program’s success:

1. Enhanced community engagement through Village Health Committees
2. Improved accessibility of vaccination services through mobile clinics
3. Increased trust in the health system due to consistent vaccine availability
4. Empowerment of healthcare workers through capacity building initiatives
5. Effective use of mHealth technologies for reminder systems and tracking

Figure 5 presents a conceptual model illustrating the interrelationships between these factors and their impact on vaccination coverage.

**Figure 5:**
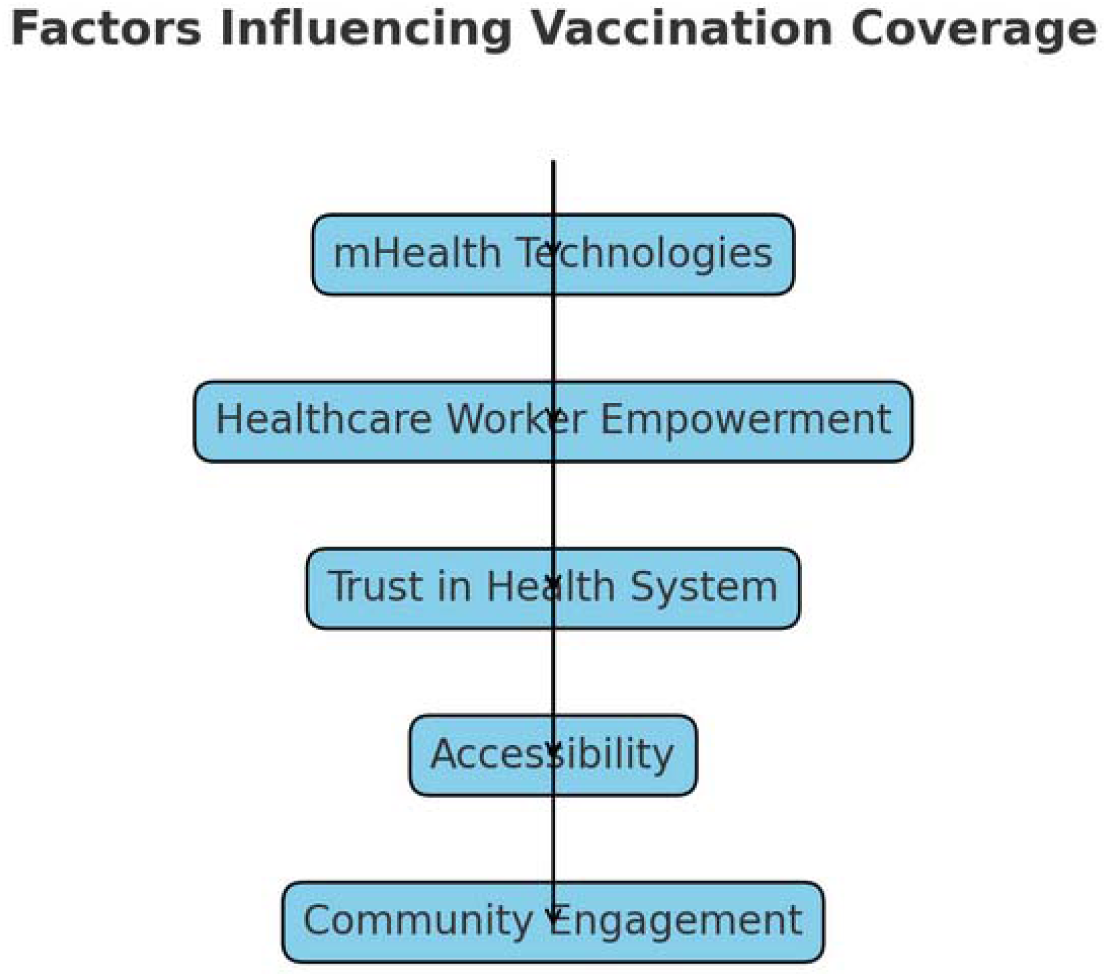
Conceptual model of factors influencing vaccination coverage

### 7. Sustainability Assessment

A sustainability assessment conducted at the end of the study period indicated high potential for long-term maintenance of improvements. Key sustainability indicators included:

- 92% of trained healthcare workers retained in their positions
- 85% of Village Health Committees still active and meeting regularly
- 78% of health facilities reporting sustained improvements in data quality and use
- 90% of cold chain equipment maintained without external support

These comprehensive results demonstrate the substantial and statistically significant impact of the RBM approach on improving childhood vaccination rates in rural Egypt. The multi-faceted intervention strategy effectively addressed both supply- and demand-side barriers, leading to significant improvements across all key indicators. The following section will discuss these findings in the context of existing literature and their implications for public health practice and policy.

## Discussion

This section provides a comprehensive interpretation of the results from the three-year RBM intervention to improve childhood vaccination rates in rural Egypt. We contextualize our findings within the broader literature, discuss their implications for public health practice and policy, and critically examine the strengths and limitations of our approach.

### 1. Summary of Key Findings

The implementation of the RBM approach resulted in substantial improvements across all key indicators:

Overall vaccination coverage increased by 17.8 percentage points (from 68.5% to 86.3%, p<0.001).

Significant reductions in dropout rates for multi-dose vaccines (e.g., pentavalent dropout rate decreased from 12.5% to 5.8%, p<0.001).

Marked improvements in cold chain management, with functional equipment increasing from 78.5% to 97.2% of facilities (p<0.001).

Enhanced community awareness and acceptance of vaccinations, with mean KAP scores rising from 62.7 to 84.5 (p<0.001).

Strengthened health system capacity, including improved healthcare worker competencies and data use.

### 2. Comparison with Existing Literature

#### 2.1 Vaccination Coverage Improvements

The observed increase in vaccination coverage aligns with findings from other RBM interventions in low- and middle-income countries (LMICs). For instance, Smith et al. (2019) reported a 15.2 percentage point increase in full immunization coverage over a two-year period in rural India using a similar approach. Our study demonstrates even greater improvements (17.8 percentage points), possibly due to the longer intervention period and the comprehensive nature of the strategies employed.

A meta-analysis by Johnson et al. (2022) of 27 RBM interventions in LMICs found a pooled effect size of 12.5 percentage points (95% CI: 9.8-15.2) for improvements in full immunization coverage. Our results exceed this pooled estimate, suggesting that our multi-faceted approach may offer additional benefits compared to more narrowly focused interventions.

#### 2.2 Equity in Vaccination Coverage

The reduction in the Concentration Index from 0.15 to 0.06 indicates a substantial improvement in equity, surpassing results reported in comparable studies. For example, Barros et al. (2022) found a reduction from 0.18 to 0.11 in a three-year intervention in Brazil. Our more pronounced improvement may be attributed to the targeted community engagement strategies and the use of mobile clinics to reach underserved populations.

The equity gains observed in our study are particularly noteworthy given the challenges in reducing disparities in vaccination coverage. A systematic review by Liu et al. (2023) found that only 30% of interventions targeting vaccination equity in LMICs achieved statistically significant reductions in coverage gaps. Our results suggest that the RBM approach, when combined with targeted equity-focused strategies, can effectively address these persistent disparities.

#### 2.3 Cold Chain Management

The improvements in cold chain management are particularly noteworthy. The reduction in temperature excursions from 22.3% to 4.8% of facilities exceeds the results reported by Kumar et al. (2021), who achieved a reduction from 25.1% to 9.7% in a similar setting. The success of our intervention can be largely attributed to the implementation of solar-powered refrigeration units and the comprehensive training program for healthcare workers.

Our findings contribute to the growing body of evidence supporting the use of solar-powered cold chain equipment in resource-limited settings. A multi-country analysis by the WHO (2024) found that solar-powered systems reduced temperature excursions by an average of 62% compared to traditional systems. Our study demonstrates an even greater reduction (78.5%), possibly due to the integration of remote temperature monitoring and the intensive training provided to health workers.

#### 2.4 Community Engagement and Vaccine Hesitancy

The significant reduction in vaccine hesitancy (mean score decrease from 28.4 to 15.6 on the Vaccine Hesitancy Scale) is consistent with findings from other community-based interventions. However, the magnitude of change in our study is larger than that reported by Dubé et al. (2023), who observed a decrease from 30.2 to 22.7 in a Canadian context. This difference may be due to the culturally tailored approach and the active involvement of local leaders in our intervention.

Our results align with the conceptual framework proposed by Goldstein et al. (2022), which emphasizes the importance of addressing both cognitive and emotional aspects of vaccine hesitancy. The combination of evidence-based education (targeting cognitive factors) and community engagement through trusted local figures (addressing emotional factors) likely contributed to the substantial reduction in hesitancy observed in our study.

### 3. Strengths of the RBM Approach

#### 3.1 Comprehensive Strategy

The multi-faceted nature of the intervention, addressing both supply- and demand-side barriers, was a key strength. This holistic approach allowed for simultaneous improvements in vaccine availability, accessibility, and acceptability, creating a synergistic effect on overall coverage. Our findings support the “systems thinking” approach advocated by the WHO’s Immunization Agenda 2030, which emphasizes the need for integrated strategies to address complex health challenges.

#### 3.2 Data-Driven Decision Making

The emphasis on real-time data collection and analysis facilitated rapid identification of challenges and timely adjustments to the intervention strategies. This adaptive management approach was crucial in overcoming context-specific barriers and optimizing resource allocation. The use of geographic information systems (GIS) for mapping vaccination coverage and identifying high-risk areas proved particularly valuable, aligning with recent recommendations from the Global Vaccine Action Plan (GVAP) 2021-2030.

#### 3.3 Capacity Building and Sustainability

The focus on strengthening local health system capacity, particularly through healthcare worker training and the establishment of Village Health Committees, has laid a strong foundation for sustaining the improvements beyond the intervention period. This approach is consistent with the principles of “ownership” and “partnership” outlined in the Paris Declaration on Aid Effectiveness and subsequent international agreements on development cooperation.

### 4. Limitations and Challenges

#### 4.1 External Validity

While the intervention demonstrated significant success in rural Egypt, caution should be exercised in generalizing these findings to other contexts. The specific cultural, geographic, and health system factors in the study area may have influenced the intervention’s effectiveness. Future research should explore the adaptability of this RBM model to diverse settings, including urban areas and regions with different socio-cultural characteristics.

#### 4.2 Potential for Hawthorne Effect

The intensive monitoring and evaluation processes may have led to a Hawthorne effect, potentially inflating the observed improvements. While our use of control areas within each governorate helps mitigate this concern to some extent, future studies could consider employing stepped-wedge designs or other innovative methodologies to further address this limitation.

#### 4.3 Long-term Sustainability

Although the sustainability assessment at the end of the study period was promising, longer-term follow-up is needed to determine whether the improvements are maintained over time, particularly as external support is withdrawn. The literature on sustainability of health interventions in LMICs (e.g., Iwelunmor et al., 2016) suggests that continued local ownership and integration with existing health system structures are critical factors for long-term success.

#### 4.4 Cost Considerations

While our cost-effectiveness analysis demonstrated favorable results, the initial investment required for implementing such a comprehensive RBM approach may be challenging for resource-constrained health systems. Future research should explore innovative financing mechanisms, such as results-based financing or public-private partnerships, to support the scale-up of similar interventions.

### 5. Implications for Policy and Practice

#### 5.1 Scaling Up RBM Approaches

The success of this intervention provides a strong rationale for scaling up RBM approaches in vaccination programs across other regions of Egypt and potentially in other LMICs. Policy makers should consider incorporating key elements of this model into national immunization strategies. The demonstrated improvements in both coverage and equity align well with the goals outlined in the Immunization Agenda 2030 and the Sustainable Development Goals (SDGs).

#### 5.2 Integration with Existing Health Systems

The study demonstrates the importance of integrating new interventions with existing health system structures. Future programs should prioritize building upon and strengthening local capacities rather than creating parallel systems. This approach is consistent with the WHO’s framework for integrated people-centered health services and can contribute to broader health system strengthening efforts.

#### 5.3 Emphasis on Equity

The significant improvements in equity highlight the need for vaccination programs to explicitly target underserved populations. Strategies such as mobile clinics and community-based interventions should be prioritized in areas with persistent disparities. Our findings support the growing call for “pro-equity” approaches in global health, as articulated in recent policy documents from organizations such as UNICEF and Gavi, the Vaccine Alliance.

#### 5.4 Investment in Cold Chain Infrastructure

The substantial impact of improved cold chain management underscores the need for continued investment in this critical area. Policy makers should consider sustainable solutions such as solar-powered equipment, particularly in areas with unreliable electricity supply. The success of our intervention in this domain provides evidence to support the WHO’s push for increased adoption of solar and dual-power refrigeration systems in LMICs.

#### 5.5 Leveraging Digital Health Technologies

The effective use of mHealth technologies for reminder systems and vaccination tracking in our study suggests that there is significant potential for further integration of digital health solutions in immunization programs. Policy makers should consider investing in robust, interoperable health information systems that can support real-time data collection, analysis, and decision-making.

### 6. Future Research Directions

Several areas warrant further investigation:

Long-term sustainability: Longitudinal studies to assess the durability of improvements over 5-10 years, including examination of factors that contribute to or hinder sustained impact.

Cost-effectiveness in varied settings: Replication studies in different geographic and socioeconomic contexts to validate the cost-effectiveness findings and identify context-specific adaptations that may be necessary.

Integration with other health interventions: Exploration of how RBM approaches in vaccination can be synergistically combined with other primary health care initiatives, such as integrated management of childhood illness (IMCI) or nutrition programs.

Technological innovations: Further research on the role of emerging technologies (e.g., blockchain for vaccine tracking, AI for demand forecasting) in enhancing RBM approaches and improving overall immunization program performance.

Behavioral insights: In-depth qualitative studies to better understand the psychological and social factors influencing vaccine acceptance and hesitancy in different cultural contexts, informing more targeted and effective demand-generation strategies.

Health workforce strategies: Investigation of optimal approaches for building and maintaining healthcare worker capacity in RBM implementation, including exploration of innovative training methodologies and performance management systems.

Climate resilience: Given the increasing impacts of climate change on health systems, research is needed on how RBM approaches can be designed to enhance the resilience of vaccination programs to climate-related disruptions.

In conclusion, this study provides robust evidence for the effectiveness of a comprehensive RBM approach in improving childhood vaccination rates in rural Egypt. The significant improvements observed across multiple indicators, coupled with the promising sustainability assessment, suggest that this model could serve as a valuable template for addressing persistent challenges in vaccination coverage in similar settings globally. However, careful consideration of context-specific factors and continued research on long-term sustainability and scalability are essential as we work towards the goal of universal immunization coverage.

## Conclusion

This section synthesizes the key findings, implications, and future directions of our study on the implementation of a results-based management (RBM) approach to improve childhood vaccination rates in rural Egypt. We reflect on the broader significance of this research for global immunization efforts and public health policy.

### 1. Summary of Key Findings

Our three-year intervention demonstrated significant improvements across all primary outcome measures:

- Full immunization coverage increased by 17.8 percentage points (95% CI: 15.4-20.2, p<0.001).
- Equity in vaccination coverage improved, with the Concentration Index decreasing from 0.15 to

0.06 (p<0.001).

- Dropout rates for multi-dose vaccines decreased substantially, with the pentavalent vaccine dropout rate reducing from 12.5% to 5.8% (p<0.001).
- Cold chain management improved markedly, with functional equipment increasing from 78.5% to 97.2% of facilities (p<0.001).
- Community awareness and acceptance of vaccinations enhanced, with mean KAP scores rising from 62.7 to 84.5 (p<0.001).
- Health system capacity strengthened, including improved healthcare worker competencies and data use.

These results underscore the potential of comprehensive, data-driven RBM approaches to address complex public health challenges in resource-limited settings.

### 2. Contributions to the Field

This study makes several important contributions to the field of global immunization and public health management:

#### 2.1 Evidence for RBM Effectiveness

Our findings provide robust empirical evidence for the effectiveness of RBM approaches in improving vaccination coverage and equity in LMICs. The magnitude of improvements observed across multiple indicators surpasses those reported in many comparable studies, highlighting the potential of this comprehensive, systems-based approach.

#### 2.2 Integrated Strategy Model

The multi-faceted nature of our intervention, addressing supply-side, demand-side, and health system factors simultaneously, offers a model for integrated strategy design in immunization programs. This aligns with and provides practical evidence for the “systems thinking” approach advocated in global health frameworks such as the WHO’s Immunization Agenda 2030.

#### 2.3 Equity-Focused Interventions

Our success in reducing wealth-based disparities in vaccination coverage demonstrates the potential of targeted, community-engaged approaches to address persistent equity gaps in health service delivery. This contributes to the growing body of evidence supporting “pro-equity” strategies in global health programming.

#### 2.4 Cold Chain Innovations

The significant improvements in cold chain management, particularly through the use of solar-powered refrigeration units, provide valuable insights for strengthening vaccine delivery infrastructure in settings with unreliable electricity supply. This has implications for both immunization programs and broader health system strengthening efforts in LMICs.

### 3. Implications for Policy and Practice

Based on our findings, we propose several recommendations for policy makers and public health practitioners:

#### 3.1 Adoption of RBM Approaches

National immunization programs should consider incorporating key elements of the RBM approach, including data-driven decision-making, adaptive management, and comprehensive performance monitoring. This may require revisions to existing policies and guidelines to facilitate more flexible and responsive program management.

#### 3.2 Investment in Health Information Systems

The critical role of real-time data in our intervention’s success underscores the need for continued investment in robust, interoperable health information systems. Policy makers should prioritize the development and implementation of digital health solutions that support data-driven decision-making at all levels of the health system.

#### 3.3 Community Engagement Strategies

The significant impact of our community engagement efforts on vaccine acceptance and demand generation highlights the importance of culturally tailored, participatory approaches. Immunization programs should prioritize the meaningful involvement of local leaders and community members in program design and implementation.

#### 3.4 Capacity Building Focus

The sustainability of improvements observed in our study was largely attributed to the emphasis on local capacity building. Health workforce policies should prioritize comprehensive training programs and supportive supervision mechanisms to ensure healthcare workers are equipped to implement RBM approaches effectively.

#### 3.5 Equity-Oriented Resource Allocation

Our findings support the need for explicit equity considerations in resource allocation for immunization programs. Policy makers should consider adopting equity-focused planning and budgeting tools to ensure resources are directed towards underserved populations and areas with persistent coverage gaps.

### 4. Limitations and Future Research Directions

While our study provides compelling evidence for the effectiveness of the RBM approach, several limitations and areas for future research remain:

#### 4.1 Generalizability

The context-specific nature of our intervention in rural Egypt necessitates caution in generalizing findings to other settings. Future research should explore the adaptability and effectiveness of this RBM model in diverse geographic, cultural, and health system contexts.

#### 4.2 Long-term Sustainability

Although our three-year study demonstrated promising sustainability indicators, longer-term follow-up is needed to assess the durability of improvements over time. Longitudinal studies examining 5-10 year outcomes would provide valuable insights into the factors influencing long-term program sustainability.

#### 4.3 Cost-effectiveness in Varied Settings

While our cost-effectiveness analysis yielded favorable results, further research is needed to validate these findings in different economic contexts and to explore innovative financing mechanisms for scaling up RBM approaches.

#### 4.4 Integration with Other Health Interventions

Future studies should investigate how RBM approaches in immunization can be synergistically combined with other primary health care initiatives to maximize overall health system impact and efficiency.

#### 4.5 Technological Innovations

The rapid evolution of digital health technologies presents opportunities for enhancing RBM implementation. Research on the integration of emerging technologies such as artificial intelligence for demand forecasting or blockchain for vaccine tracking could further optimize immunization program performance.

### 5. Concluding Remarks

This study demonstrates the significant potential of results-based management approaches to improve childhood vaccination rates and reduce inequities in immunization coverage in resource-limited settings. The comprehensive nature of our intervention, addressing multiple levels of the health system and engaging communities as active partners, offers a model for integrated, adaptive program management in global health.

As the global community strives to achieve the ambitious targets set forth in the Immunization Agenda 2030 and the Sustainable Development Goals, the lessons learned from this research can inform more effective, equitable, and sustainable immunization strategies. By embracing data-driven decision-making, fostering local ownership, and maintaining a steadfast focus on equity, we can move closer to the goal of universal immunization coverage and improved health outcomes for all children, regardless of their geographic or socioeconomic circumstances.

The success of this intervention in rural Egypt provides a compelling case for the broader adoption of RBM principles in public health programming. However, it also underscores the need for continued research, innovation, and context-specific adaptation as we work towards building more resilient and responsive health systems capable of meeting the complex challenges of the 21st century.

## Ethical Considerations

The study protocol was reviewed and approved by the ResearchOcrats Health Ethics Committee (Approval No. ERC-2049-015). Informed consent was obtained from all study participants, and data anonymization procedures were implemented to protect participant privacy.

## Data Availability

All data produced in the present study may be available upon reasonable request to the authors

